# *Plasmodium vivax* malaria serological exposure markers: assessing the degree and implications of cross-reactivity with *P. knowlesi*

**DOI:** 10.1101/2021.12.09.21266664

**Authors:** Rhea J Longley, Matthew J Grigg, Kael Schoffer, Thomas Obadia, Stephanie Hyslop, Kim A. Piera, Narimane Nekkab, Ramin Mazhari, Eizo Takashima, Takafumi Tsuboi, Matthias Harbers, Kevin Tetteh, Chris Drakeley, Chetan E. Chitnis, Julie Healer, Wai-Hong Tham, Jetsumon Sattabongkot, Michael T White, Daniel J Cooper, Giri S Rajahram, Bridget E. Barber, Timothy William, Nicholas M Anstey, Ivo Mueller

## Abstract

Serological exposure markers are a promising tool for surveillance and targeted interventions for *Plasmodium vivax* malaria. *P. vivax* is closely related to the zoonotic parasite *P. knowlesi*, which also infects humans. *P. vivax* and *P. knowlesi* are co-endemic across much of South East Asia, making it important to design *P. vivax* serological markers that minimise cross-reactivity in this region. Our objective was to determine the degree of IgG antibody cross-reactivity against a panel of *P. vivax* serological markers in samples from human participants with *P. knowlesi* malaria. We observed higher levels of IgG antibody reactivity against *P. vivax* proteins that had high levels of sequence identity with their *P. knowlesi* ortholog. IgG reactivity peaked at 7 days post *P. knowlesi* infection and was short-lived, with minimal responses detected at 1-year post-infection. Using these data, we designed a panel of 8 *P. vivax* proteins with low-levels of cross-reactivity with *P. knowlesi*. This panel was able to accurately classify recent *P. vivax* infections whilst reducing misclassification of recent *P. knowlesi* infections.

## Introduction

In order to accelerate towards malaria elimination, new tools and interventions are needed. Malaria is caused by the parasite *Plasmodium,* with the majority of disease in humans caused by *P. falciparum, P. vivax, P. malariae, P. ovale,* and *P. knowlesi. P. falciparum* and *P. vivax* account for the largest burdens of disease. In many co-endemic regions outside Africa, infection with *P. vivax* has become the predominant cause of malaria as the overall level of transmission has declined (reviewed in^1^). Thus, whilst standard malaria control tools (such as those targeting the mosquito vector and case management) have had a significant impact on reducing transmission of *P. falciparum*, they have not had the same level of effect on *P. vivax.* This is most likely due to a number of distinct biological features of *P. vivax* infections that promote transmission. These include a cryptic endosplenic life-cycle leading to a large hidden splenic reservoir of *P. vivax* parasites^2, 3^ which sustains a high prevalence of low-density asymptomatic blood stage infections^4^ that are still capable of onward transmission^5^. Furthermore, an additional life-cycle stage in the liver, known as the hypnozoite, can remain arrested or dormant in the liver for months to years following the initial mosquito-bite induced infection, before being reactivated by currently unknown signals to re-join the life-cycle. Hepatic hypnozoites are major reservoirs for transmission in communities, with up to 80% of detected blood-stage infections attributed to hypnozoite relapse rather than new mosquito bite-induced infections^6–8^.

To address the unique challenge to malaria elimination posed by *P. vivax*, we have developed a novel surveillance tool: *P. vivax* serological exposure markers. By measuring total IgG antibody responses to a carefully selected panel of 8 *P. vivax* proteins, we can classify individuals as recently exposed to a blood-stage *P. vivax* infection within the past 9-months with 80% sensitivity and 80% specificity^9^. Due to the frequency of *P. vivax* relapses^10^, we hypothesize our *P. vivax* serological exposure markers can be used to indirectly identify hypnozoite carriers – individuals who are more likely to go on to develop recurrent *P. vivax* infections. Our serological markers can thus be used as an effective surveillance tool for identifying clusters of *P. vivax* infections for efficient targeting of resources^11^, and for designing public health interventions relying on “serological testing and treatment” (seroTAT)^12^. SeroTAT can be used to identify hypnozoite carriers who can then be treated with appropriate anti-malarial drugs (including an anti-liver stage component such as primaquine or tafenoquine) as part of an elimination campaign^9^.

The major species of *Plasmodium* causing disease in humans have different geographic distributions^13^ and different levels of relatedness^14^. It is therefore important to characterise potential cross-reactivity against our *P. vivax* serological exposure markers in individuals who have had recent exposure to other *Plasmodium* infections. We identified no patterns of cross-reactivity with recent *P. falciparum* exposure in our original cohort studies^9^, with the caveat that few individuals had *P. falciparum* infections in those observational cohorts (in Thailand, Brazil and the Solomon Islands). In the limited number of prior studies assessing *P. vivax* – *P. falciparum* cross-reactive antibody responses^15^, mixed results were observed with both species-specific^16^ and cross-reactive antibodies detected^17, 18^. However, even if cross-reactivity between our *P. vivax* proteins in individuals with recent *P. falciparum* infections becomes evident, this is not necessarily of programmatic concern. Similar risk-factors for exposure in co-endemic regions are present and there is potential benefit in treating individuals with *P. falciparum* infections with anti-hypnozoite drugs^19^ (due to the high risk of *P. vivax* recurrence following *P. falciparum* infections^20–22^). *P. vivax* is more closely related to the zoonotic parasite *P. knowlesi* than *P. falciparum*^14^ and serological cross-reactivity has been reported^23–25^. *P. vivax* and *P. knowlesi* are co-endemic in humans across much of South East Asia^26^. Risk-factors for exposure to *P. knowlesi* and *P. vivax* also differ^27^, as do spatial clusters of infections^25^, treatment^28^ and risk mitigation strategies^27^. For these reasons, and to optimise the utility of serological markers in South East Asia, it is of particular importance to characterise cross-reactivity against the *P. vivax* serological exposure markers in individuals with recent *P. knowlesi* infections.

Here, we aimed to assess cross-reactivity against a panel of *P. vivax* serological exposure markers (including our previously identified top combination of 8 markers^9^) in samples from two clinical cohorts of patients with confirmed *P. knowlesi* malaria. We assessed whether our algorithm would classify these *P. knowlesi* patients as *P. vivax*-exposed or not, and whether we could improve our serological markers for use in *P. vivax-P. knowlesi* co-endemic areas by selecting proteins with absent or lower levels of cross-reactivity.

## Results

### Comparison of *P. vivax* and *P. knowlesi* protein sequences

We hypothesized that potential antibody cross-reactivity could relate to the level of sequence identity and thus we first constructed a pipeline to identify and then compare the sequences of our *P. vivax* serological exposure markers with their identified *P. knowlesi* orthologs. Two different analytical approaches were used: 1) by accessing the list of orthologs on PlasmoDB for each *P. vivax* protein and 2) through a NCBI BlastP search of the protein sequence construct. *P. knowlesi* orthologs were found for 17 of 21 *P. vivax* proteins via PlasmoDB (using either the original macaque H strain^29^ or the later human red blood cell adapted A1H1 line^30^) and for 20/21 by NCBI BlastP (any strain) (Table 1). Using both methods each of the 21 *P. vivax* proteins had at least one identified *P. knowlesi* ortholog. When multiple orthologs were found, the top hit was selected by highest identity. The similarity and identity values for the hits on PlasmoDB were obtained by aligning the full-length protein sequences provided in the database using the EMBOSS Needle protein alignment tool; the identity values for the NCBI BlastP hits were obtained from the BlastP search using only the protein construct sequence. The sequence identity of the full-length protein sequences (search strategy 1) ranged from 11-19% for MSP7F (A1H1 line or H strain, respectively) to 83.4% for MSP8. A similar trend was found using the specific protein construct and NCBI BlastP (search strategy 2) however the percent identity was often higher using this method (Table 1).

**Table 1:** Sequence comparison of the P. vivax proteins with their P. knowlesi orthologs. Full-length protein sequences were first compared using the orthologs listed in the PlasmoDB database, then the protein construct sequences were compared using NCBI BlastP to identify the top hits (see Table S1 for the amino acid region of each construct). NA indicates there were no matches for the database search. Proteins are ordered by highest percentage sequence identity using the NCBI BlastP method. The H strain and A1H1 line gave identical results except for MSP7F; the H strain results are listed, A1H1 gave 11.5% identity and 18.8% similarity. * Indicates top 8 P. vivax protein serological exposure marker.

| *P. vivax* |  | *P. knowlesi* |  |  |  |  |  |  |
| --- | --- | --- | --- | --- | --- | --- | --- | --- |
|  |  | PlasmoDB (H or A1H1 strain) |  |  |  |  | NCBI (any strain) |  |
| *P. vivax* Protein | PlasmoDB code | # Hits | Top Hit | Syteny | Similarity (%) | Identity (%) | Top Hit | Identity (%) |
| MSP8 * | PVX_097625 | 1 | PKNH_1031500 | Y | 91.2 | 83.4 | AFL93300.1 | 84.52 |
| Pv-fam-a* | PVX_112670 | 24 | PKNH_1300800 | Y | 72.8 | 65.8 | OTN66803.1 | 83.11 |
| MSP1-19* | PVX_099980 | 1 | PKNH_0728900 | Y | 74.4 | 64.2 | AZL87433.1 | 81.31 |
| RAMA* | PVX_087885 | 0 | NA | NA | NA | NA | OTN68698.1 | 73.61 |
| RIPR | PVX_095055 | 1 | PKNH_0817000 | Y | 82.9 | 71.9 | XP_002258810.1 | 73.43 |
| Pv-fam-a* | PVX_096995 | 24 | PKNH_0200900 | Y | 50.9 | 41.1 | OTN66147.1 | 70.28 |
| PTEX150 | PVX_084720 | 1 | PKNH_0422900 | Y | 83.9 | 73.2 | OTN66840.1 | 72.02 |
| DBPII Sal 1 | PVX_110810 | 3 | PKNH_0623500 | Y | 64.7 | 52.1 | OTN68355.1 | 68.72 |
| DBPII AH | AAY34130.1 | 3 | PKNH_0623500 | Y | 64.7 | 51.9 | OTN68355.1 | 68.21 |
| Sexual stage antigen s16 | PVX_000930 | 1 | PKNH_0304300 | Y | 78.6 | 67.1 | OTN68464.1 | 65.45 |
| TRAP | PVX_082735 | 1 | PKNH_1265400 | Y | 81.4 | 69.1 | AAG24613.1 | 64.23 |
| MSP3b | PVX_097680 | 15 | PKNH_1457400 | N | 42 | 28.6 | OTN63878.1 | 62.5 |
| MSP5 | PVX_003770 | 2 | PKNH_0414200 | Y | 65.4 | 49.9 | AAT77929.1 | 46.26 |
| Hypothetical | PVX_097715 | 0 | NA | NA | NA | NA | ANP24393.1 | 40.65 |
| EBP region II* | KMZ83376.1 | 0 | NA | NA | NA | NA | QPL17772.1 | 35 |
| RBP2b<sub>1986-2653</sub> | PVX_094255 | 2 | PKNH_0700200 | N | 52.2 | 29.1 | OTN67427.1 | 34.53 |
| MSP7F | PVX_082670 | 15 | PKNH_0726500 | N | 30.1 | 19 | AND94835.1 | 27.53 |
| RBP2b<sub>161-1454</sub>* | PVX_094255 | 2 | PKNH_0700200 | N | 52.2 | 29.1 | OTN67427.1 | 26.1 |
| RBP2a | PVX_121920 | 2 | PKNH_0700200 | N | 46.7 | 25.4 | OTN67427.1 | 25.77 |
| MSP7L | PVX_082700 | 0 | NA | NA | NA | NA | AND94835.1 | 24.29 |
| MSP3a* | PVX_097720 | 15 | PKNH_1457400 | N | 45.8 | 31.6 | NA | NA |

### *P. vivax* protein-specific IgG antibody response in *P. knowlesi* clinical case samples

To assess potential cross-reactivity against the *P. vivax* proteins we measured longitudinal total IgG antibody responses in patients with PCR-confirmed *P. knowlesi* monoinfection, enrolled in two clinical trials, denoted ACTKNOW (recruited over 2012-2014^31^) and PACKNOW (recruited over 2016-2018^32^) (Table 2). The two sample sets were initially analysed separately given the risk of prior *P. vivax* infection was lower at the time of the PACKNOW trial^33^. There were no differences in demographic variables such as age, sex and self-reported history of malaria infections between the patients in the two trials (Table 2).

**Table 2:** Characteristics of patients in two P. knowlesi clinical trial cohorts used in the current study.

|  | **ACTKNOW (n=99)** | **PACKNOW (n=41)** |
| --- | --- | --- |
| PCR-confirmed *Plasmodium* spp. | *P. knowlesi* | *P. knowlesi* |
| Collection year | 2012-2014 | 2016-2018 |
| Age (years), median (range)<sup>a</sup> | 34 (3-78) | 36 (20-70) |
| Sex, number female<sup>b</sup> (percent) | 29 (29.3%) | 6 (14.6%) |
| Self-reported fever days, median (range)<sup>a</sup> | 5 (0-14) | 4 (3-14) |
| Self-reported previous malaria infection, number yes<sup>b</sup> (percent) | 34 (34.3%) | 17 (41.5%) |
| Parasitaemia (parasites/μl), median (range)<sup>a</sup> | 2857 (56-43,721) | 2690 (37-185,553) |
| Treatment administered | Randomised to artesunate-mefloquine (n=52) or chloroquine (n=47) | Artemether-lumefantrine |
| *Serology details* |  |  |
| Sample size at enrolment | 99 | 41 |
| Follow-up timepoints | 3: day 0, 7, 28 | 5: day 0, 7, 14, 28, 365 |
| Total # samples assessed | 296<sup>c</sup> | 166<sup>d</sup> |

<sup>a</sup>Statistical difference between ACTKNOW and PACKNOW cohorts assessed by Mann-Whitney test, not significant. <sup>b</sup>Statistical difference between ACTKNOW and PACKNOW cohorts assessed by Fisher's exact test, not significant. <sup>c</sup>One sample missing serology data at day 0.
<sup>d</sup>Samples were not available at all time-points for all patients.

We first assessed potential cross-reactivity against the *P. vivax* proteins using 99 patients from the ACTKNOW *P. knowlesi* clinical trial cohort, with plasma samples available at the time of diagnosis of *P. knowlesi* infection (day 0) and follow-up at day 7 and day 28. Median parasitemia prior to treatment (day 0) was 2,857 parasites/µL (IQR 648-7,177), with all subjects negative for asexual stage parasites and 95% negative for sexual stages by the day 7 timepoint^31^. Antibody responses were compared to malaria-naïve negative control samples from the non-endemic cities of Melbourne, Australia, Bangkok, Thailand and Rio, Brazil (n=369), with a protein-specific seropositivity cut-off defined as the average of the negative controls plus two times the standard deviation. We observed an increase in the median IgG antibody response against all *P. vivax* proteins at day 7 compared to day 0, with IgG levels declining from the peak response by day 28 (Fig. 1). For 17 of 21 *P. vivax* proteins, median IgG levels at day 7 (the peak of the response) were above the sero-positivity cut-off. Of those 17 proteins, for five the median IgG level had dropped below the sero-positivity cut-off by day 28. For all *P. vivax* proteins there were at least some patients with responses above the sero-positivity cut-off at all three time-points (see Supplementary Figure 1 for the change in IgG levels at the individual level over time, n=98 patients with matched data from all three time-points). We explored whether acquisition of cross-reactive IgG antibodies at day 0 was related to delayed presentation to the clinic (and therefore delayed enrolment into the study), using duration of fever as a surrogate for delayed presentation/duration of infection. There was no correlation between day 0 IgG antibodies and duration of fever for 19/21 *P. vivax* proteins; there was a weak negative correlation for both DBPII constructs (Spearman correlation, AH r=-0.25, Sal1 r=-0.2, p<0.05 see Figure S2). The highest levels of potential cross-reactivity were observed for MSP1-19, MSP8, Pv-fam-a (PVX_096995) and RAMA, with median IgG levels at day 7 greater than 10-fold compared to the seropositivity cut-off. All 4 of these proteins are used in the classification algorithm based on a combination of antibody responses to 8 proteins, with MSP1-19 and RAMA particularly having an important contribution to the classification performance based on a variable importance plot^9^.

**Fig. 1.**
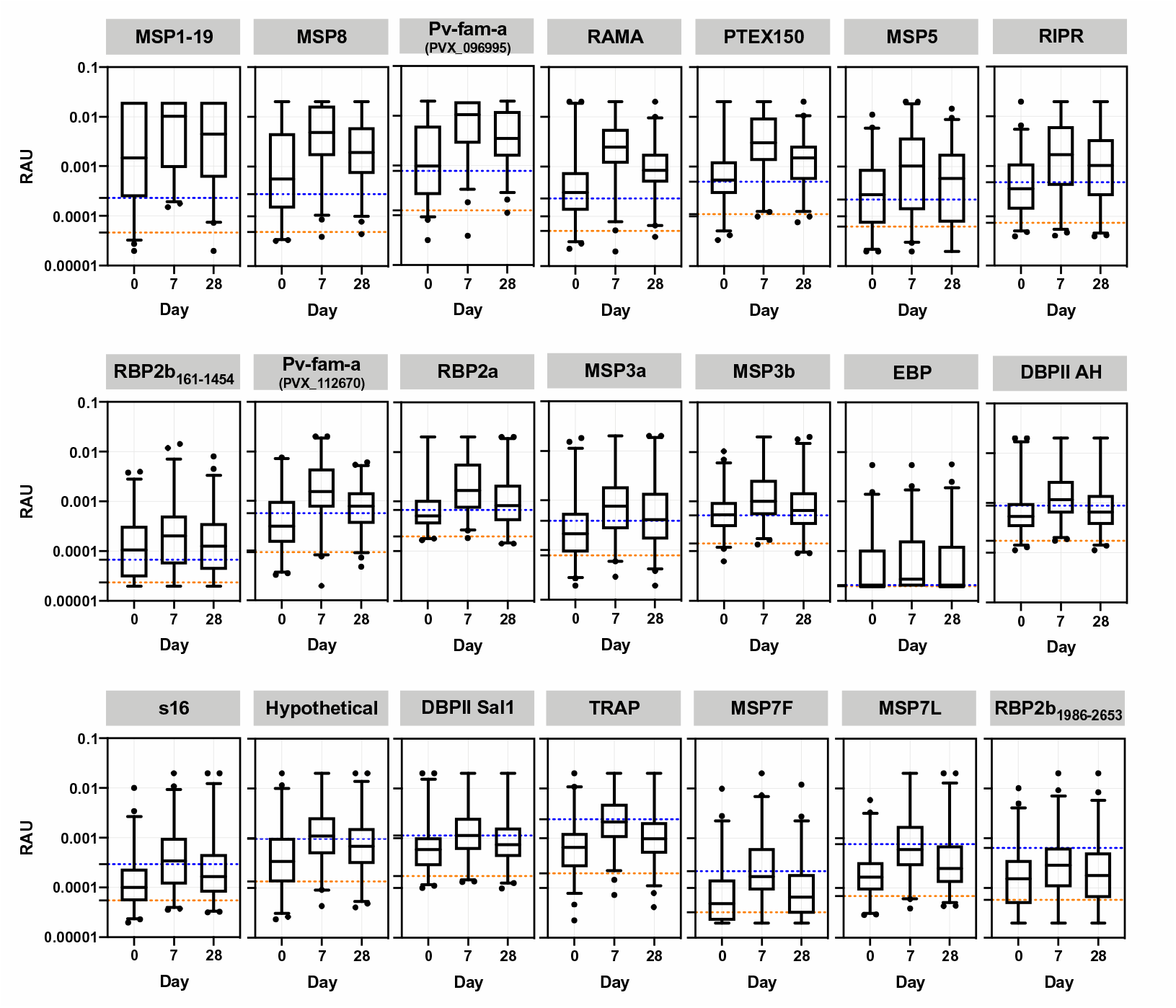
IgG antibody levels against 21 *P. vivax* proteins in patients with clinical *P. knowlesi* infections. IgG levels were measured against the 21 *P. vivax* proteins using a multiplexed antibody assay. Individual patients (n=99) had longitudinal samples obtained and run at the time of diagnosis of *P. knowlesi* infection (day 0), and days 7 and 28 following enrolment (ACTKNOW cohort). Day 0 has data from 98 samples due to the wrong sample being re-run following quality control. Results are expressed as the relative antibody units (RAU). Proteins are ordered by highest level of median IgG at day 7 compared to the seropositivity cut-off. Dashed lines indicate the malaria-naïve negative control samples: orange = average of the negative control samples; blue = seropositivity cut-off (average plus 2x standard deviation).

In order to confirm the results of the ACTKNOW study and to determine the longevity of the cross-reactive antibody response over 1 year (median 369 days; IQR 365-394) post *P. knowlesi*-infection, we measured IgG antibody levels against the 21 *P. vivax* proteins in the PACKNOW clinical trial cohort samples (n=41 patients with up to 5 time-points available, recruited over 2016-2018, at a time of near-elimination of *P. vivax* in Sabah). These patients were all treated with artemether-lumefantrine. Median parasitemia prior to treatment (day 0) was 2,690 parasites/ µL (IQR 572-14,317), with all subjects negative for asexual stage parasites by the day 7 timepoint^32^. There was no significant difference in parasitaemia at day 0 between the ACTKNOW and PACKNOW subjects (median 2857 vs 2690 respectively, see Table 2). Similar to the ACTKNOW cohort, we observed a peak in the total IgG response to all 21 *P. vivax* proteins at days 4-15 post enrolment, with responses generally declining by the day 27-30 time-point (Fig. 2). Total IgG antibodies against the protein *P. vivax* MSP1-19 were an exception, with elevated levels maintained at the day 27-30 time-point and also at 1-year. By 1-year after *P. knowlesi* infection, the median *P. vivax* total IgG antibodies had declined to below the sero-positivity cut-off for all other proteins (Fig. 2). There were fewer *P. vivax* proteins (n=11/21) with a peak median total IgG response above the sero-positivity cut-off in the PACKNOW samples (collected across 2016-2018) compared to the ACTKNOW samples (collected across 2012-2014) (n=17/21), although this was not statistically significant (Fisher’s exact test, p=0.1). There was a significantly higher magnitude total IgG response to 9/21 *P. vivax* proteins in the ACTKNOW compared to PACKNOW cohorts at day 7 (Mann-Whitney U test, see Figure S3). The top 4 *P. vivax* proteins with the highest levels of cross-reactivity evident were the same as the ACTKNOW cohort, being MSP1-19, MSP8, Pv-fam-a (PVX_096995) and RAMA (median IgG levels at day 7 more than 6-fold higher than the sero-positivity cut-off).

**Figure 2:**
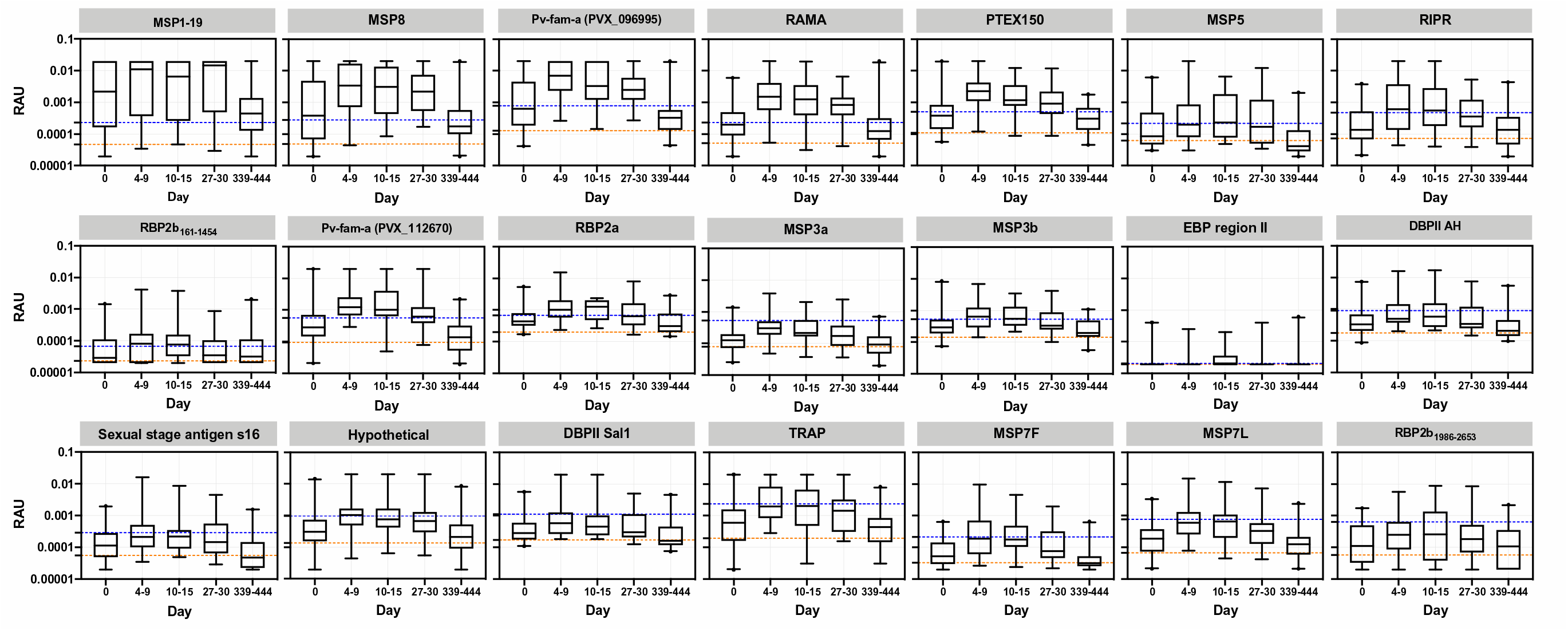
IgG antibody levels against 21 *P. vivax* proteins in patients up to 1 year post clinical *P. knowlesi* infections. IgG levels were measured against the 21 *P. vivax* proteins using a multiplexed antibody assay. Samples were obtained and run at the time of *P. knowlesi* infection (day 0) (n=41), days 4-9 (n=35), days 10-15 (n=15), days 27-30 (n=33) and days 339-444 (n=42) following enrolment (PACKNOW cohort). Results are expressed as the relative antibody units (RAU). Proteins are ordered as per Figure 1. Dashed lines indicate the malaria-naïve negative control samples: orange = average of the negative control samples; blue = seropositivity cut-off (average plus 2x standard deviation).

There was a statistically significant correlation between the fold change comparing the median IgG level at the peak at day 7 with the seropositivity cut-off and the percentage sequence identity of the *P. knowlesi* and *P. vivax* orthologs for both the ACTKNOW and PACKNOW cohorts (Fig. 3A-B) (Spearman correlation, r=0.63, p=0.0023 and r=0.56, p=0.0083, respectively) and using median IgG antibody data from both patient cohorts combined (r=0.69 p=0.0006, Fig. 3C).

**Fig. 3.**
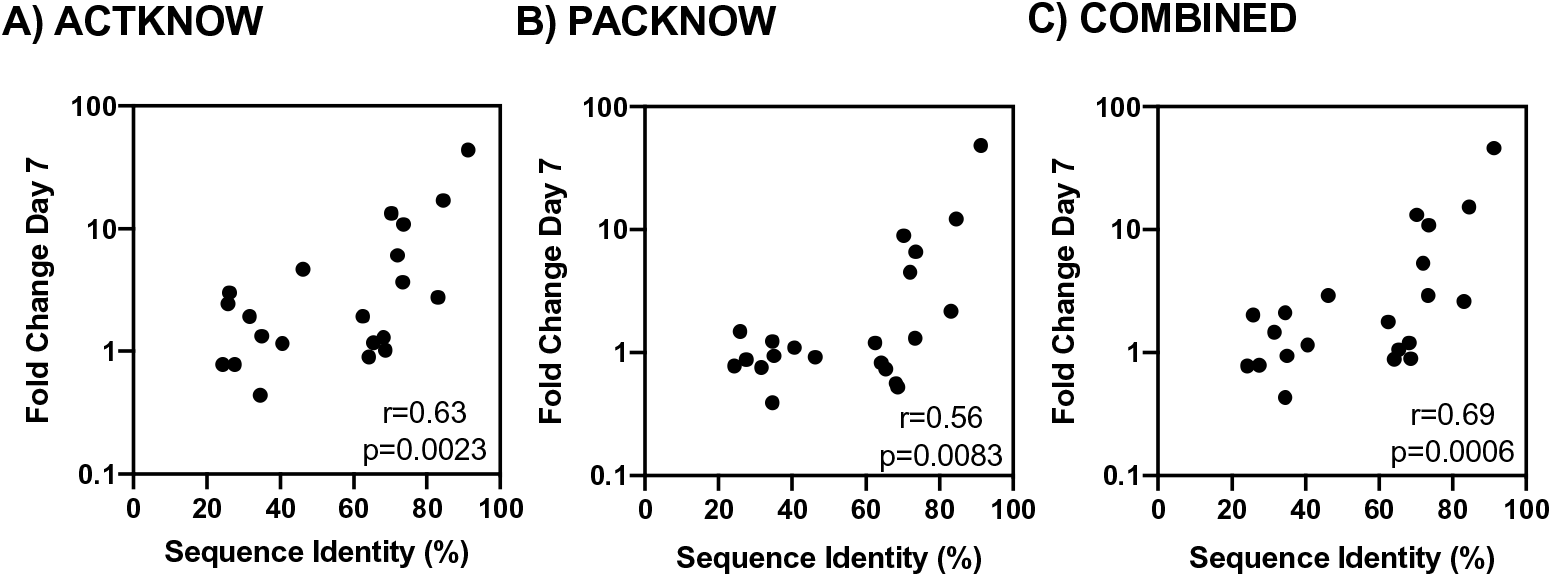
Correlation between the peak anti-*P. vivax* IgG level at day 7 and the percentage sequence similarity of the *P. vivax* and *P. knowlesi* orthologs. The median IgG level at day 7 (the peak of the response) was divided by the seropositivity cut-off to generate the fold change at the peak compared to the background. The percentage sequence identity was calculated for the protein construct sequence (not full-length for all proteins) using NCBI BlastP. Where there was no hit for NCBI BlastP the PlasmoDB method was used (see Table 1). A Spearman’s correlation was performed to determine the relationship of the fold change with the sequence identity using data from all 21 *P. vivax* proteins, for the A) ACTKNOW r=0.63 p=0.0023, B) PACKNOW cohorts r=0.56 p=0.0083 and C) ACTKNOW and PACKNOW combined (median antibody level of n=134 *P. knowlesi* patients at Day 7 divided by the seropositivity cut-off) r=0.69 p=0.0006.

### Classification of recent exposure to *P. vivax* blood-stage infections in the *P. knowlesi* cohorts

As high levels of cross-reactivity (>6-10 fold increase over the sero-positivity cut-off) were evident between four of our top 8 *P. vivax* proteins in the two series of *P. knowlesi* clinical case samples (MSP1-19, MSP8, Pv-fam-a (PVX_096995), RAMA), we next used these data in our previously developed classification algorithm to determine whether any of the samples from patients with current or recent *P. knowlesi* infections were classified as recently exposed to *P. vivax.* The top 8 *P. vivax* proteins used in the classification algorithm are: MSP1-19, MSP8, Pv-fam-a (PVX_096995), RAMA, RBP2b_161-1454_, Pv-fam-a (PVX_112670), MSP3a, and EBP. We utilised a balanced sensitivity and specificity target of 79%. We ran the algorithm on both the ACTKNOW and PACKNOW cohorts at all time-points for which we had data, and the proportion of patients classified as positive by the algorithm varied between 17-82% (Table 3). The trend was for the highest numbers of *P. knowlesi* patients to be classified as recently exposed to *P. vivax* at the day 7 timepoint in both cohorts (82% ACTKNOW, 77% PACKNOW), in line with the highest peak IgG activity. The lowest proportion of *P. knowlesi* patients classified as positive was for the PACKNOW cohort at Day 365 (7/42, 17%).

**Table 3:**
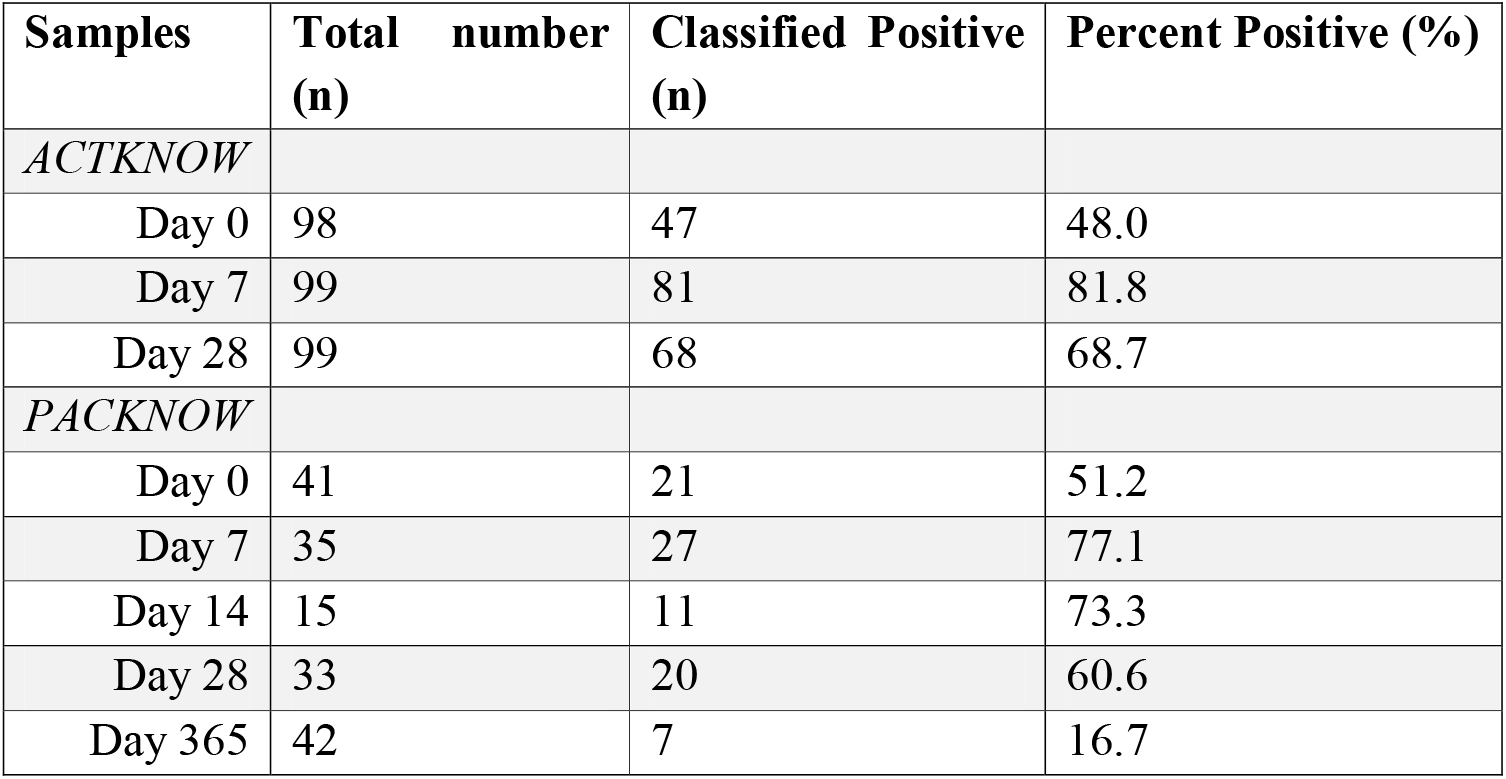
Output from *P. vivax* classifier using IgG antibody responses from top 8 *P. vivax* serological exposure markers. A balanced 79% specificity, 79% sensitivity target was used.

| Samples | Total number (n) | Classified Positive (n) | Percent Positive (%) |
| --- | --- | --- | --- |
| *ACTKNOW* |  |  |  |
| Day 0 | 98 | 47 | 48.0 |
| Day 7 | 99 | 81 | 81.8 |
| Day 28 | 99 | 68 | 68.7 |
| *PACKNOW* |  |  |  |
| Day 0 | 41 | 21 | 51.2 |
| Day 7 | 35 | 27 | 77.1 |
| Day 14 | 15 | 11 | 73.3 |
| Day 28 | 33 | 20 | 60.6 |
| Day 365 | 42 | 7 | 16.7 |

For the ACTKNOW cohort, of the 99 patients, 46 were classified positive by the algorithm at all three time-points. 20 were classified positive at day 7 and day 28, 14 at day 7 only, two at day 28 only and one at day 0 and day 7. 16 patients were not classified as positive at any timepoint. Whilst there was variability in the available plasma samples post enrolment for the PACKNOW cohort, 9 patients were not classified positive at any timepoint and all those who were positive at a later timepoint (day 28 or day 365) were previously positive for at least one of the earlier timepoints (i.e. day 0, 7 or 14). The exceptions were 1 patient who was positive at only day 28 and 1 patient positive at only day 365 (the latter had notably high antibody levels at day 365, reaching equivalent to a 1/50 dilution of the positive control pool against 4 of 8 antigens).

### Design and application of an adjusted *P. vivax*-specific serological marker panel

In both the ACTKNOW and PACKNOW *P. knowlesi* clinical cohorts, more than half the patients at day 7 and day 28 post-infection were classified as recently exposed to *P. vivax* blood-stage parasites using our existing algorithm with our set of 8 *P. vivax* serological exposure markers. This indicated our current serological exposure marker panel was not *P. vivax*-specific. We hypothesized that using proteins with low-levels of cross-reactivity in the *P. knowlesi* infected patients would reduce misclassification. We thus designed a new panel of 8 *P. vivax* serological exposure markers by selecting the proteins with the lowest levels of cross-reactivity in the *P. knowlesi* clinical samples (as determined by the median IgG level at Day 7 from the ACTKNOW sample set compared to the background, as described for Fig. 3). These proteins were: RBP2b_1986-2653_, MSP7L, MSP7F, TRAP, DBPII Sal1, hypothetical (PVX_097715), sexual stage antigen s16, and EBP (all with a fold change of less than 2). DBPII AH was not selected given this is a variant of the DBP region II that was already included (Sal1 strain included). We re-trained the algorithm on our existing Thai, Brazilian and Solomon Islands datasets^9^ for these 8 *P. vivax* proteins. Youden’s J was used to determine the best trade-off balancing sensitivity and specificity, assuming equal importance of false positives and false negatives in the classification process. The algorithm trained with the new panel of 8 *P. vivax* proteins achieved 72.4% sensitivity and 70.8% specificity respectively, for classifying recent *P. vivax* infections within the prior 9-months (Fig. 4A). When applied to confirmed *P. knowlesi* clinical cases, we identified a reduced number of 210/462 (45%) samples being classed as recently exposed to *P. vivax* (Table 4) (compared to 281/462 (61%) using the original panel). Misclassification was still high at day 7 post-*P. knowlesi* infection (69.7% ACTKNOW and 80% PACKNOW) but less than 58% at day 28 and 7% at day 365. The greatest reduction in numbers and proportions of *P. knowlesi* patients classified positive by the algorithm was for the ACTKNOW cohort.

**Fig. 4.**
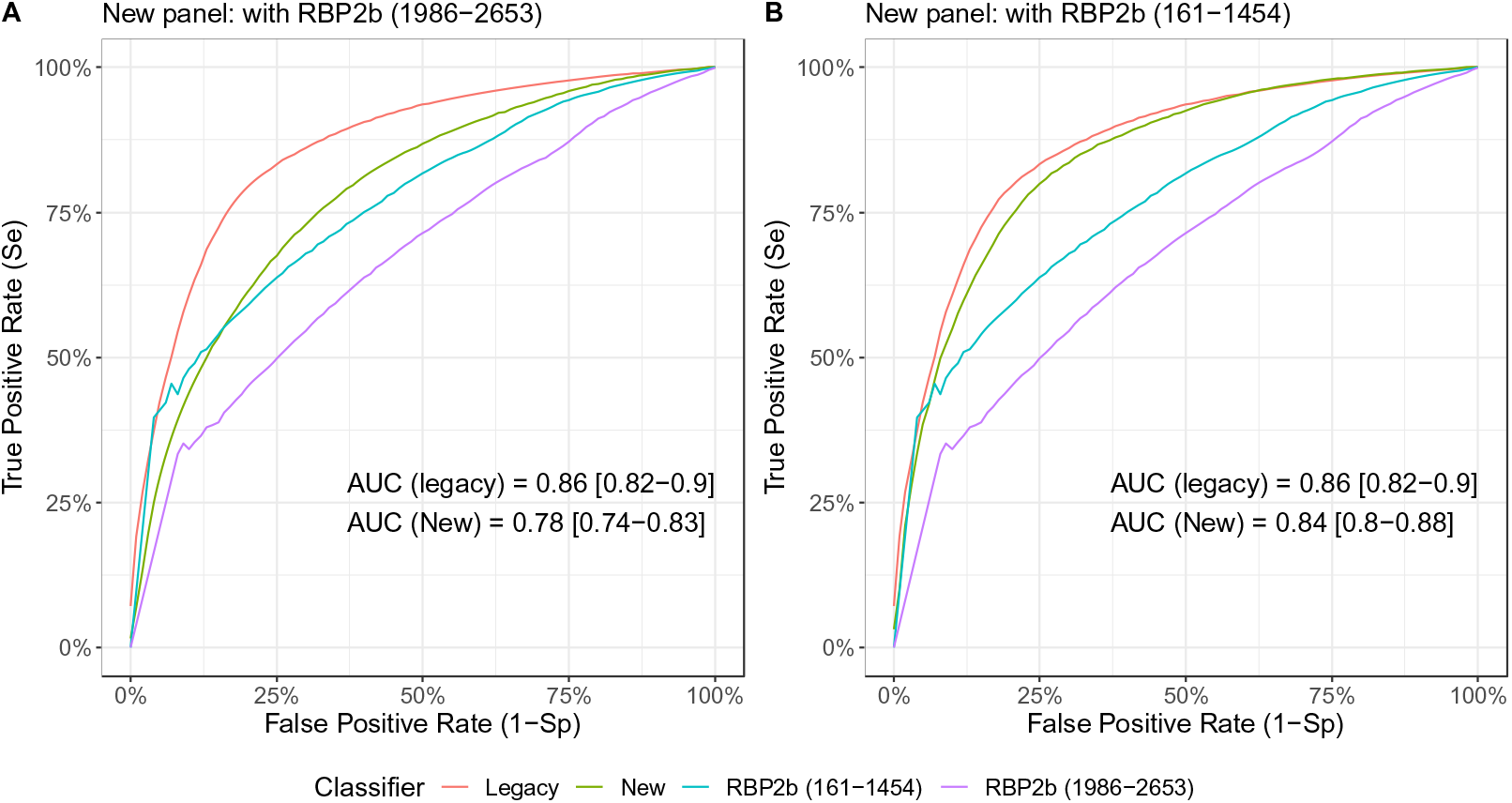
Classification of recent *P. vivax* infections using an adjusted *P. vivax* antigen panel. Receiver operator characteristic curve for classifying individuals with PCR-detected *P. vivax* infections within the prior 9 months, using a random forest classification algorithm with antibody responses to a modified set of 8 *P. vivax* antigens (green, “new”) compared to the original 8 (red, “legacy”). In panel A) RBP2b_1986-2653_, MSP7L, MSP7F, TRAP, DBPII Sal1, hypothetical (PVX_097715), s16 and EBP, and B) RBP2b_161-1454_, MSP7L, MSP7F, TRAP, DBPII Sal1, hypothetical (PVX_097715), s16 and EBP. Both panels A and B also include classification using either of the RBP2b_1986-2653_ (purple) or RBP2b_161-1454_ (blue) responses alone.

**Table 4:**
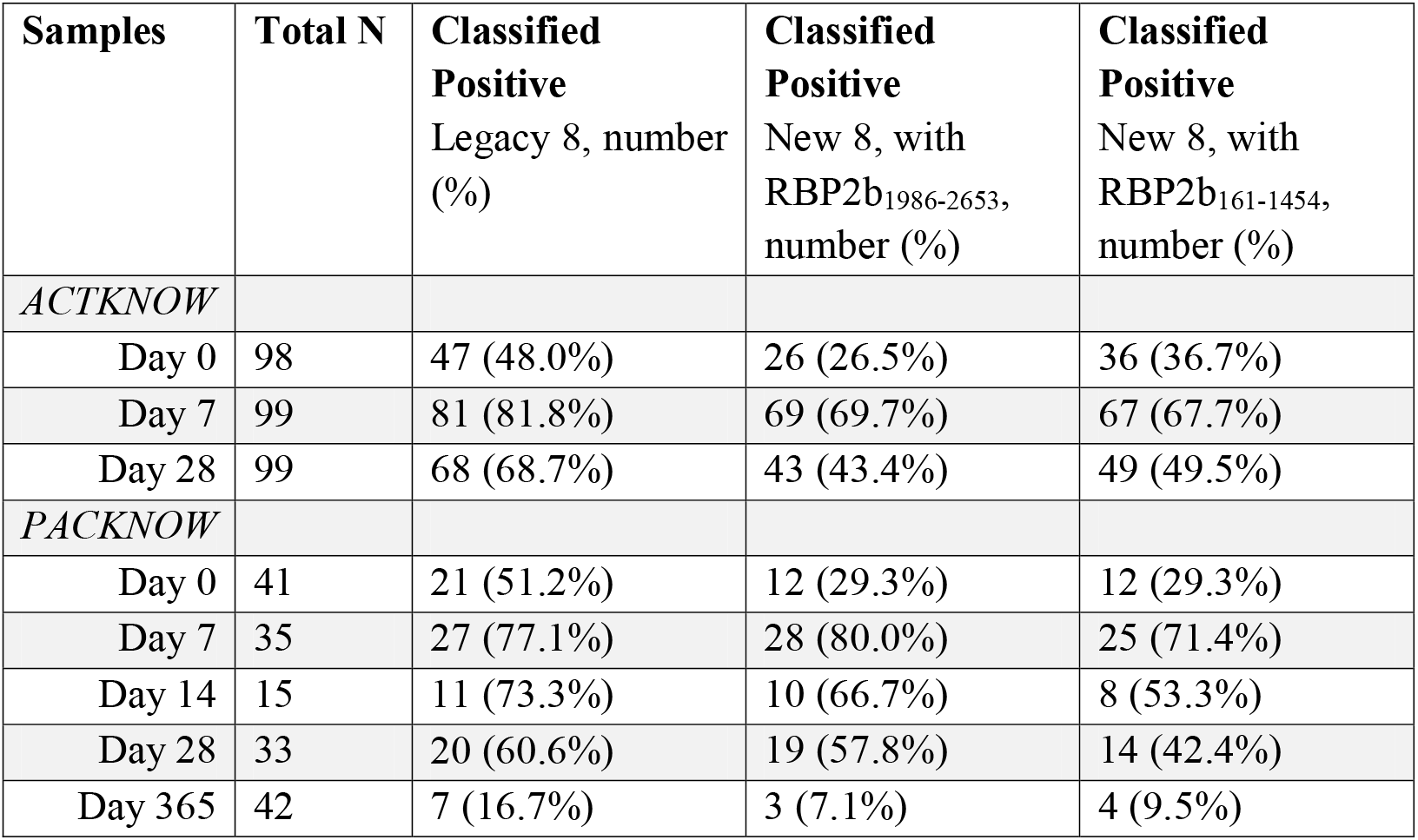
Output from *P. vivax* classifier using IgG antibody responses from two adjusted panels of 8 *P. vivax* serological exposure markers with low levels of *P. knowlesi*-cross-reactivity. A classification of previous exposure was taken when predicted probability was greater than a cut off corresponding to the respective sensitivity and specificity targets of 72.4% and 70.8% (RBP2b_1986-2653_) or 78.7% and 76.9% (RBP2b_161-1454_). The results from Table 2 are included for comparison.

Whilst the new combination of 8 *P. vivax* proteins reduced overall misclassification of *P. knowlesi* clinical cases, the sensitivity and specificity for correctly classifying recent *P. vivax* infections (72.4%, 70.8%) was substantially lower than the original 8 (80% for both^9^). We hypothesized this was because the new panel did not include the top classifier RBP2b_161-1454_ (see Fig. 4. and Longley, White et al., 2020^9^). As this protein also had relatively low-levels of cross-reactivity in the *P. knowlesi* samples at day 7 (fold change 2.9 compared to the seropositivity cut-off), we replaced RBP2b_1986-2653_ in the new panel with RBP2b_161-1454_. This improved classification of recent *P. vivax* infections in the Thai, Brazilian and Solomon Islands datasets to 78.7% sensitivity and 76.9% specificity (Fig. 4B), which is more comparable to the original classifier. Applying this trained classification algorithm to the *P. knowlesi* sample sets resulted in 215/462 (47%) samples being misclassified (Table 4), which is less than when using the original algorithm (281) and similar to the 210 misclassified when using RBP2b_1986-2653_ in the new panel (new algorithm RBP2b_1986-2653_ vs RBP2b_161-1454_ P=0.79, Fisher’s exact test). At day 7 post-*P. knowlesi* infection 68-71% were classified as recently exposed to *P. vivax*, reducing to 42-50% at day 28 and 10% at day 365. Those patients positive at day 365 were also classified positive at earlier timepoints.

### Association of peak antibody levels with age and *P. knowlesi* parasitaemia

Antibody levels are commonly associated with age (i.e. increasing antibodies with increasing age, due to both being a proxy for exposure and age itself), and sometimes with the antigenic input as measured by blood-stage parasitaemia. We further explored whether the cross-reactive antibodies identified at day 7 were associated with either age or *P. knowlesi* parasitaemia (as recorded at enrolment) through regression analyses. This analysis was performed using peak antibody levels at day 7 from both the ACTKNOW and PACKNOW cohorts (n=134). There was a weak positive association with age (range 3-78 years) for 11/21 *P. vivax* antigens (Table 5).

**Table 5:** Associations between peak IgG anti-*P. vivax* antibody levels at day 7 in the both *P. knowlesi* cohorts combined with age. Sample size = 134. Regression analyses were performed univariably and adjusted with parasitaemia. Antigens are ordered by the fold change in the peak antibody level at day 7 in the ACTKNOW cohort compared to the seropositivity cut-off based on the negative control samples (=Fold Δ IgG). CI = confidence interval.

| Protein | Fold Δ IgG | Age (unadjusted) |  | Age (adjusted) |  |
| --- | --- | --- | --- | --- | --- |
|  |  | Coefficient (95% CI) | *p* value | Coefficient (95% CI) | *p* value |
| MSP1-19 | 43.98 | 0.012 (0.0052-0.02) | 0.001 | 0.012 (0.0052-0.020) | 0.001 |
| MSP8 | 17.07 | 0.005 (-0.0012-0.011) | 0.11 | 0.005 (-0.0012-0.011) | 0.111 |
| Pv-fam-a (PVX_096995) | 13.39 | 0.0047 (-0.00062-0.01) | 0.083 | 0.0047 (-0.00064-0.01) | 0.084 |
| RAMA | 10.86 | 0.0046 (-0.0021-0.011) | 0.175 | 0.0046 (-0.0021-0.011) | 0.175 |
| PTEX150 | 6.08 | 0.013 (0.0053-0.020) | 0.001 | 0.013 (0.0053-0.02) | 0.001 |
| MSP5 | 4.70 | 0.021 (0.013-0.029) | <0.0001 | 0.021 (0.013-0.029) | <0.0001 |
| RIPR | 3.66 | 0.014 (0.0073-0.021) | <0.0001 | 0.014 (0.0073-0.0211) | <0.0001 |
| RBP2b<sub>161-1454</sub> | 3.00 | 0.0077 (0.0012-0.014) | 0.02 | 0.0077 (0.0011-0.014) | 0.021 |
| Pv-fam-a (PVX_112670) | 2.76 | -0.0048 (-0.01-0.0036) | 0.068 | -0.0048 (-0.01-0.00038) | 0.069 |
| RBP2a | 2.45 | 0.012 (0.0071-0.018) | <0.0001 | 0.012 (0.007-0.018) | <0.0001 |
| MSP3b | 1.93 | 0.0064 (0.0014-0.011) | 0.013 | 0.0064 (0.0013-0.012) | 0.014 |
| MSP3a | 1.92 | 0.015 (0.0088-0.021) | <0.0001 | 0.015 (0.0088-0.021) | <0.0001 |
| EBP | 1.33 | 0.012 (0.0072-0.016) | <0.0001 | 0.012 (0.0072-0.016) | <0.0001 |
| DBPII AH | 1.31 | 0.0044 (-0.00086-0.0097) | 0.1 | 0.0044 (-0.0008-0.0096) | 0.099 |
| S16 | 1.18 | 0.0061 (0.00020-0.012) | 0.043 | 0.0061 (0.00018-0.012) | 0.044 |
| Hypothetical | 1.16 | 0.0038 (-0.0028-0.010) | 0.258 | 0.0038 (-0.0028-0.01) | 0.26 |
| DBPII Sal1 | 1.02 | 0.0051 (-0.001-0.011) | 0.102 | 0.0051 (-0.001-0.011) | 0.102 |
| TRAP | 0.90 | 0.007 (0.0014-0.013) | 0.015 | 0.007 (0.014-0.013) | 0.015 |
| MSP7L | 0.78 | 0.0057 (-0.00012-0.012) | 0.055 | 0.0057 (-0.00013-0.0115) | 0.055 |
| MSP7F | 0.78 | 0.0066 (-0.0029-0.013) | 0.06 | 0.0066 (-0.0025-0.013) | 0.059 |
| RBP2b<sub>1986-2653</sub> | 0.44 | 0.0042 (-0.0063-0.012) | 0.266 | 0.0042 (-0.0033-0.118) | 0.272 |

In prior studies of *P. knowlesi* malaria age is associated with parasitaemia^34, 35^, thus we also adjusted the age analyses for parasitaemia and the findings remained the same (Table 5). Parasitaemia ranged from 56-185,553 parasites/µL, and there was no association between *P. knowlesi* parasitaemia and peak antibody levels at day 7 for 19/21 *P. vivax* antigens (Table S2). For DBPII AH there was a negative association between parasitaemia and antibody levels, whereas for MSP7F there was a positive association between parasitaemia and antibody levels. There were also higher antibody levels in females compared to males for four proteins (MSP5, MSP3b, MSP3a, DBPII AH) (Table S2); three of these also had a significant positive association with age which remained after adjustment for sex. No pattern was observed between these associations with the ranking of antigens by cross-reactivity (Table 5).

## Discussion

*P. vivax* serological markers, that reflect recent exposure to blood-stage *P. vivax* parasites and can indirectly identify hypnozoite carriers, could play an important role in accelerating malaria elimination programs. In this study, we aimed to identify whether cross-reactive antibodies against our *P. vivax* serological markers were present in patients with recent *P. knowlesi* infections. Using serial plasma samples from two cohorts of patients with *P. knowlesi* malaria, we found that cross-reactive antibodies are induced against some of the *P. vivax* proteins assessed. A statistically significant association was seen with the level of sequence identity between the *P. vivax* and *P. knowlesi* orthologs. The peak cross-reactive antibody response was at day 7 following infection, which is the same timing as the peak *P. knowlesi*-antibody response reported previously using a similar sample set from Sabah, Malaysia^24^. This is also in line with the timing of peak IgG antibody responses reported for other species such as *P. falciparum* and *P. vivax* (within the first two weeks post-treatment)^36–38^. Antibodies against the *P. vivax* proteins had reduced in magnitude by day 28 post-infection, and the median IgG levels were below the seropositivity cut-off for all but one *P. vivax* protein (MSP1-19) at 1-year post-*P. knowlesi* infection.

The two cohorts of *P. knowlesi* clinical case samples used here were obtained during different time periods, 2012-2014 and 2016-2018. The incidence of *P. vivax* infections in Sabah, Malaysia in 2012-2014 was modest and transmission had declined even further by 2016-2018^33^. Therefore, the risk that the *P. knowlesi*-infected patients had recent past exposure to *P. vivax* was low in both cohorts (notably, 14 patients were retrospectively excluded from the ACTKNOW study due to *P. vivax* infections detected by PCR whilst only 1 was excluded from the PACKNOW study, see Methods). The number of *P. vivax* proteins with antibody levels above the sero-positive cut-off at the peak of the response was higher in the ACTKNOW cohort in 2012-2014 at 80.95% (17/21) compared to 52.38% (11/21) in the PACKNOW cohort in 2016-2018 (this difference was not statistically significant). There was also a significantly higher magnitude of the IgG response in the ACTKNOW cohort compared to the PACKNOW cohort for 9/21 *P. vivax* proteins at day 7. There was no significant difference in the age between the two cohorts nor the parasitaemia at enrolment (Table 2). Overall, there were still anti-*P. vivax* antibody responses present in the later PACKNOW cohort and thus our interpretation of these responses as cross-reactive is by far the most likely explanation given the evidence (rather than recent past exposure to *P. vivax* infections). Furthermore, the *P. vivax* protein RBP2b is the top serological exposure marker and highly accurate at predicting recent *P. vivax* infections even when used alone^9^. The lack of antibody responses detected against RBP2b in the *P. knowlesi*-clinical cohorts suggests it is unlikely these patients also had recent *P. vivax* infections.

Our findings are in line with, but extend upon, past research. An extensive study of *P. vivax* –*P. knowlesi* antibody cross-reactivity to 19 blood-stage proteins was recently published^23^, which included four of the proteins we assessed in the current study (MSP1, MSP8, DBP, RAMA). The authors generated rabbit antibodies against the 19 *P. vivax* proteins and found they recognised *P. knowlesi* blood-stage parasites through immunofluorescence assays, and were able to inhibit invasion of *P. knowlesi* into erythrocytes *in vitro*^23^. Naturally acquired antibodies from individuals living in endemic areas were also analysed, with evidence of cross-reactive antibodies from either of *P. vivax* or *P. knowlesi* patients against *P. vivax* or *P. knowlesi* recombinant proteins (building upon the authors’ prior work on one *P. vivax* protein, apical asparagine rich protein^39^). Of relevance to our study, they found significant levels of reactivity in *P. knowlesi* patient sera against *P. vivax* MSP8^23^, which is supported by our current data. The 19 *P. vivax* proteins assessed by Muh and colleagues had high levels of sequence identity with their *P. knowlesi* orthologs (>58.2%)^23^, with the exception of two RBP proteins (1a and 1b, 22.3 and 26.9%, respectively), and this could explain the universal cross-reactivity they identified. In contrast, our panel of *P. vivax* proteins had a large spread of sequence identity with their *P. knowlesi* orthologs, with 9 proteins having less than 50% sequence identity, which enabled us to demonstrate the positive relationship between sequence identity and antibody cross-reactivity. A lower level of sequence identity comes with a decrease in the likelihood of continuous linear epitopes, which is in support of our finding. Further work would need to be undertaken to determine how lower sequence identity affects discontinuous conformational epitopes. Overall, we found the presence and/or level of cross-reactivity is antigen-specific. Our work further extends upon existing findings by demonstrating that cross-reactive antibody responses in *P. knowlesi* patients against *P. vivax* proteins are short-lived.

The most cross-reactive protein in our panel was MSP1-19 (>40-fold increase at day 7 compared to sero-positivity cut-off). This magnitude of response is equivalent to that reached against a *P. knowlesi*-specific protein (*Pk*SERA antigen 3) in a subset of these same *P. knowlesi* patient samples (50-fold at day 7 compared to day 0)^24^. Previous assessment of antibody responses against MSP1-19 in *P. falciparum, P. vivax, P. malariae* and *P. ovale* (but notably did not include assessment of *P. knowlesi*) indicated a highly species-specific response^16^. However, high-levels of cross-reactivity for MSP1-19 between *P. vivax* and *P. knowlesi* (>81% sequence identity between the Sal1 and H strains, respectively) have been reported in other studies in-line with our work^23, 24^. *P. vivax* MSP1-19 was a vaccine candidate^40^, though has not progressed to clinical trials^41^. There remains interest in this candidate^42, 43^, and the high levels of cross-reactivity in our study with *P. knowlesi* suggest that potential cross-species immunity could be possible^15^. Recent work has used *P. knowlesi* as a model for screening inhibitory activity of *P. vivax* antibodies^44^. Polyclonal antibodies against the *P. vivax* proteins DBP, two 6-cysteine proteins (Pv12, Pv14) and the GPI-anchored micronemal antigen (GAMA) were able to inhibit invasion of wild-type *P. knowlesi*, demonstrating that cross-species functional immunity is possible (at least *in vitro*). No inhibition was evident against wild-type *P. knowlesi* when using antibodies against *P. vivax* MSP3a and MSP7L, which is consistent with our findings of minimal cross-reactivity in *P. knowlesi* patients for these proteins. In addition to MSP1-19, we observed high levels of potential cross-reactive antibody responses against the *P. vivax* proteins MSP8, Pv-fam-a (PVX_096995) and RAMA. These antigens could thus be considered for assessment of growth inhibitory activity in the *P. knowlesi* model^44^ and considered as potential targets for cross-species protection. This is an important consideration given the currently limited landscape of vaccine development specifically for *P. knowlesi*^45^. In some previously co-endemic areas of South East Asia, the rise in incidence of symptomatic *P. knowlesi* malaria has followed the decline in *P. vivax* malaria, supporting the idea for development of a *P. vivax* vaccine that provides cross-reactive immunity against *P. knowlesi*^45^. To-date there has been limited assessment of these three *P. vivax* proteins as vaccine candidates. Unfortunately, a number of the most promising *P. vivax* vaccine candidates (including RBP2b, EBP, MSP3a^46^) have low sequence identify with their *P. knowlesi* orthologs and limited evidence of cross-reactivity in our study. Whilst not an objective of our current study, we did identify a number of interesting associations between infection related variables and antibodies against *P. vivax* DBPII, including shorter fever duration and lower parasitemia in *P. knowlesi* patients with higher antibodies. This suggests some effect of pre-existing immunity leading to a better ability to control the infection, thus warrants further attention.

All four of the most highly cross-reactive proteins are within our top 8 panel of *P. vivax* serological exposure markers^9^. However, levels of antibodies against our top serological marker, RBP2b (N and C terminal constructs), were low. When we applied our previously trained classification algorithm to the current *P. knowlesi* clinical data sets, 77-82% of *P. knowlesi* patients at the peak antibody time-point (Day 7) in each cohort were classified as recently exposed to *P. vivax* blood-stage parasites in the past 9-months. This reduced to 61-69% at Day 28 and 17% at Day 365. As mentioned, at the time of the PACKNOW cohort (2016-2018) local *P. vivax* transmission in Sabah, Malaysia had approached elimination and nearly all malaria was from *P. knowlesi*^33^. Thus, in this cohort there is less risk that the anti-*P. vivax* antibody responses measured are due to past exposure to *P. vivax*, and a lower proportion of *P. knowlesi* patients were accordingly classified as recently exposed compared to the earlier ACTKNOW cohort. However, 77% of PACKNOW patients at Day 7 were classified as recently exposed to *P. vivax*, and thus the substantial cross-reactivity identified against four of our top eight proteins is resulting in this (likely) misclassification. It is important to consider this level of misclassification in areas endemic for both *P. vivax* and *P. knowlesi* in the context of how the serological exposure markers will be applied.

Multiple use-cases for *P. vivax* serological markers exist^12^. For an intervention, such as seroTAT, even with potential misclassification due to recent exposure to *P. knowlesi*, the serological markers would still outperform mass drug administration in terms of specificity. For surveillance purposes, such as identifying clusters of infections or levels of residual transmission to better target the limited resources of malaria control programs, it will be important to define whether the recent exposure was due to *P. vivax* or *P. knowlesi.* We therefore assessed performance of an adjusted panel of 8 *P. vivax* serological markers, which were selected due to low-levels of cross-reactivity in the *P. knowlesi* clinical cases. Suboptimal performance was initially observed, but after substituting one fragment of RBP2b_1986-2653_ for the top performing RBP2b_161-1454_, this new panel of 8 markers were still able to classify recent *P. vivax* infections with 78.7% sensitivity and 76.9% specificity, whilst reducing the misclassification of recent *P. knowlesi* infections to 68-71% at the peak day 7 timepoint. By day 28 this was <50% and importantly at one year post *P. knowlesi* infection, 10%. This provides proof of principle that an adjusted panel of serological markers could be applied for use in co-endemic areas, but further optimisation would be needed to completely reduce the risk of misclassifying recent *P. knowlesi* infections. This residual cross-reactivity at one year may assume greater importance for sero-surveillance purposes, as the proportion of all *Plasmodium* infections due to *P. knowlesi* rises with *P. vivax* elimination strategies in co-endemic areas^45^. It would be useful to have plasma samples collected between 1-12 months post *P. knowlesi*-infection to better understand the dynamics of the cross-reactive antibody response. An alternative approach would be to include a panel of *P. knowlesi-*specific markers into the *P. vivax* serological assay, which could be cross-referenced to inform on recent *P. knowlesi* exposure. Recently a panel of recombinant *P. knowlesi* proteins has been developed for use as serological tools with care being taken in their design to avoid regions of sequence with high levels of identity with other *Plasmodium* species^24^. Whilst further validation of species-specificity is required, at least one protein (*P. knowlesi* serine repeat antigen 3 antigen 2) was able to identify *P. knowlesi* exposed individuals^24^. Further work could also assess the use of other Ig isotypes or IgG subclasses detected against the *P. vivax* serological exposure markers, instead of total IgG, as another avenue for reducing misclassification at later time-points post *P. knowlesi*-exposure given the possible variability in their longevity^36, 47^. It may also be possible to design smaller *P. vivax* protein fragments of the top markers that have very low sequence identitiy with their *P. knowlesi* orthologs.

Our study provides important evidence of antibody cross-reactivity against *P. vivax* proteins in patients with *P. knowlesi* infections, that are relatively short-lived, and provides an approach for limiting the potential misclassification of *Plasmodium* species by serology in co-endemic areas. This data will be useful when interpreting *P. vivax* sero-surveillance results in settings co-endemic for *P. knowlesi.* It will be important to expand upon our findings to assess potential cross-reactivity with other zoonotic^48^ and non-zoonotic *Plasmodium* species, even though for the latter the levels are expected to be lower due to the further divergence in genetic relatedness with other species^14^. Further assessment of recently developed *P. knowlesi* markers of exposure^24^ in *P. vivax* endemic areas is warranted to confirm limited cross-reactivity for those specific proteins between the two species. For our findings to be applied, the new panel of 8 *P. vivax* proteins with limited evidence of cross-reactivity with *P. knowlesi* needs to be validated for a) the ability to identify individuals at risk of recurrent *P. vivax* infections and b) the ability to identify *P. vivax* hypnozoite carriers.

## Materials and Methods

### Ethical approvals

Samples from two clinical trials conducted in Sabah, Malaysia, were used for this study (ACTKNOW^31^ and PACKNOW^32^). Ethical approvals for these studies were obtained from the Human Research Ethics Committee at the Menzies School of Health Research, Darwin, Australia (approval numbers 2012-1815 and 16-2544) and the Medical Research and Ethics Committee of the Ministry of Health Malaysia, Malaysia (approval numbers NMRR-12-89-11005 and NMRR-16-29088). All patients gave written informed consent or an attending relative gave informed consent. Ethical approval was provided by the Human Research Ethics Committee at the Walter and Eliza Hall Institute of Medical Research for use of the Malaysian and negative control samples in Melbourne (approval number 14/02).

### Study samples

Longitudinal plasma samples were collected from patients enrolled in two *P. knowlesi* clinical trials conducted in Sabah, Malaysia, denoted as ACTKNOW and PACKNOW, and four panels of malaria-naïve negative controls.

The ACTKNOW study recruited patients over 2012-2014, with the aim of comparing artesunate-mefloquine versus chloroquine for the treatment of acute uncomplicated *P. knowlesi* malaria^31^ (clinicaltrials.gov registration number NCT01708876). Patients were recruited from three hospitals (Kudat, Kota Marudu, Pitas) and were eligible for inclusion if they had microscopic diagnosis of *P. knowlesi* monoinfection, were more than 1 year of age, more than 10kg, had a negative *P. falciparum* rapid diagnostic test, and had a fever (≤ 37.5°C) or history of fever in the past 48 hours. Patients who were not confirmed by PCR to have *P. knowlesi* monoinfection were retrospectively excluded^31^. Of those excluded, 14 were due to *P. vivax* infections detected upon PCR. The duration of self-reported history of fever (days) was recorded. Patients were treated as previously described^31^, with approximately half randomised to receive chloroquine and half to receive artemisinin-based combination therapy. Plasma samples were available for the current study from 99 patients at day 0 (time of enrolment), day 7 and day 28. Data is presented from 98 samples at day 0 for 20/21 *P. vivax* proteins due to one sample failing quality control of the antibody data, and the incorrect sample being repeated by mistake.

The PACKNOW study recruited patients over 2016-2018, with the aim or comparing regularly dosed paracetamol versus no paracetamol on renal function in *P. knowlesi* malaria ^32^ (clinicaltrials.gov registration number NCT03056391). All patients received antimalarial drug treatment with artemether-lumefantrine. Patients were recruited from four hospitals (Queen Elizabeth Hospital, Keningau, Ranau, Kota Marudu) and were eligible for inclusion if they had a microscopic diagnosis of *P. knowlesi* monoinfection, fever (≥ 38°C) on admission or during the preceding 48 hours, and were more than 5 years of age. Patients were enrolled within 18 hours of commencing antimalarial treatment^32^. Patients who were not confirmed by PCR to have *P. knowlesi* monoinfection were retrospectively excluded, using a validated PCR assay^49^. In contrast to the ACTKNOW study, this only included 1 excluded due to *P. vivax* infection detected by PCR (unpublished data). Plasma samples were available for the current study from 41 patients at day 0 (time of enrolment), 7, 14, 28 and 365 (these exact time-points were not always available, exact days post-enrolment are stated in the results and relevant figure legends).

For both the ACTKNOW and PACKNOW cohorts, all patients were PCR-negative for *P. vivax* co-infection at enrolment, however it was unknown whether the *P. knowlesi* patients had past exposure to *P. vivax* parasites. *P. vivax* has been endemic in Sabah, Malaysia, but the number of malaria cases due to *P. vivax* has substantially declined from 2009 – 2017 (whilst the number of cases due to *P. knowlesi* has risen)^33^. In line, the number of patients retrospectively excluded from these studies due to *P. vivax* infection detected by PCR was 14 in ACTKNOW and only 1 in PACKNOW. All patient data variables are provided in additional file 1.

We utilised four panels of malaria-naïve negative control plasma samples as previously described^9^. Briefly, this included 102 individuals from the Volunteer Blood Donor Registry in Melbourne, Australia; 100 individuals from the Australian Red Cross, Melbourne, Australia; 72 individuals from the Thai Red Cross, Bangkok, Thailand; and 96 individuals from the Rio de Janeiro State Blood Bank, Rio de Janeiro, Brazil.

### *P. vivax* proteins

21 *P. vivax* proteins were selected for assessment of antibody responses due to their ability to classify individuals as recently exposed to *P. vivax* parasites within the prior 9-months, as previously described^9^. These included eight *P. vivax* proteins that, together, can classify recent *P. vivax* infections with 80% sensitivity and specificity. Details on the 21 *P. vivax* proteins, including the sequence construct and expression system, are provided in Table S1. In this manuscript we have referred to proteins by their annotation, with the PlasmoDB (plasmodb.org/) code in brackets if necessary. Notations for MSP7 were from Garzón-Ospina 2016^50^ and MSP3 from Kuamsab 2020^51^.

Sequence comparisons between the *P. vivax* proteins with their *P. knowlesi* orthologs was performed by identifying the orthologs in PlasmoDB or through NCBI BlastP. Only H strain and A1H1 line orthologs were selected using PlasmoDB whereas NCBI BlastP included all *P. knowlesi* strains. Sequence identity and similarity percentages were obtained using the EMBOSS Needle protein alignment tool (https://www.ebi.ac.uk/Tools/psa/emboss_needle/)^52^ or NCBI Blast (https://blast.ncbi.nlm.nih.gov/Blast.cgi) directly. PlasmoDB comparisons were made between full-length protein sequences whereas NCBI Blast used the protein sequences of expression constructs as the query sequence.

### Total IgG antibody assay

We used a multiplexed assay based on Luminex xMAP technology to measure total IgG antibody responses to the 21 *P. vivax* proteins. This assay was run on a MAGPIX instrument, as previously described^53^. Briefly, the 21 *P. vivax* proteins were coupled to individual sets of internally labelled magnetic COOH beads at previously determined optimal concentrations (see Table S1) following standard methods^53^. Antigen-specific total IgG was detected in plasma samples by incubating 500 coupled beads of each antigen per well with plasma diluted at 1:100 in PBT (1x PBS with 1% BSA and 0.05% Tween-20), followed after incubation by the addition of 0.5mg/ml PE-conjugated Donkey F(ab)2 anti-human IgG (JIR 709-116-098). On each plate, a two-fold serial dilution from 1/50 to 1/25,600 of a positive control hyperimmune plasma pool (generated from adults from PNG, a non-knowlesi-endemic region) was included. At least 15 beads per region were then acquired and read on a MAGPIX instrument as per the manufacturer’s instructions, with results expressed as the mean fluorescent intensity (MFI). All serology data is available in additional files 2 and 3.

Existing datasets of total IgG antibodies against the same *P. vivax* proteins in three observational cohort studies (in *P. vivax-*endemic areas) were also used^9^. These data were generated as previously described^9^ using a similar multiplexed assay, but with non-magnetic beads and run on a Luminex-200 instrument. Briefly, yearlong cohort studies were conducted in Thailand (Kanchanaburi and Ratchaburi provinces), and in non-knowlesi endemic Brazil (Manaus) and the Solomon Islands (Ngella) across 2013-2014. Each site enrolled between 928-1274 individuals with blood samples taken every month for qPCR detection of *P. vivax* infections and plasma stored for antibody measurements. This enabled total IgG antibody responses measured at the final visit of the yearlong cohorts to be related to the time since prior *P. vivax* infection. These existing datasets are available here: https://github.com/MWhite-InstitutPasteur/Pvivax_sero_dx.

### Statistical analysis

The raw MFI results were converted to relative antibody units (RAU) using protein-specific standard curve data, as previously described^53^. A five-parameter logistic function was used to obtain an equivalent dilution value (expressed as the RAU) compared to the PNG control plasma, with extrapolation one step beyond the lowest dilution resulting in a range of values from 1.95×10^-5^ to 0.02 RAU. The standard curve conversion was performed in R version 4.1.1. Spearman’s r correlations were performed to correlate sequence identity with relative cross-reactivity levels. Spearman r values <0.3 were considered weak, 0.3–0.7 moderate, and >0.7 strong correlations. Mann-Whitney U tests were used to compare antibody levels between various patient variables, or to compare antibody levels across cohorts. Fisher’s exact tests were used to compare categorical variables or outcomes. Correlations, Mann-Whitney U tests and Fisher’s exact tests were performed in Prism version 9. Linear regression analyses to assess associations between peak antibody levels with age, sex and parasitaemia were performed in Stata version 12. Antibody levels were first log10 transformed to better for a normal distribution.

Classification of recent *P. vivax* infections within the prior 9-months was performed using a Random Forest classification algorithm. The algorithm used antibody data against the top eight *P. vivax* serological exposure markers that we had previously identified^9^, and was trained with 4 existing datasets from our previous work (Thailand n=826, Brazil n=925, Solomon Islands n=754 and negative controls n=274)^9^. The top eight proteins were: MSP1-19, Pv-fam-a (PVX_096995), Pv-fam-a (PVX_112670), RAMA, MSP8, MSP3a, RBP2b_161-1454_, and EBP. A diagnostic target of 79% specificity and 79% sensitivity was selected.

Two modified sets of eight proteins were also tested in the classification algorithm, based on low cross-reactivity in the *P. knowlesi* clinical patient samples. A random forest classification algorithm was trained, using existing antibody data generated against the modified sets of eight proteins in our Thai, Brazilian and Solomon Islands studies^9^. The classification algorithm was cross-validated using 1000 randomly sampled, disjoint training and testing subsets, and the data presented in a receiver operating characteristic curve, with credible intervals corresponding to percentiles calculated among replicates. At every iteration, 2/3 of each cohort was used for training and 1/3 for testing. This newly modified and trained random forest algorithm was subsequently used to classify the samples from the *P. knowlesi* clinical cohorts as recently exposed to *P. vivax* blood-stage parasites in the prior 9-months, or not, with the optimal balanced diagnostic target that was achievable as per Youden’s J.

## Data Availability

All data produced in the present study are contained in the manuscript (with additional files containing the antibody and patient data).

## Acknowledgments

We acknowledge the efforts of the original study teams for collecting the *P. knowlesi* clinical samples. We thank Shazia Ruybal for support generating an automated quality control platform through R, based on prior work by Connie Li-Wai-Suen, which was also used for the standard curve conversion. We thank Sarah Miller for assistance with ethical approvals. We would like to thank the Director-General, Ministry of Health, Malaysia, for permission to publish this manuscript. We also acknowledge the Victorian State Government Operational Infrastructure Support and Australian Government NHMRC IRIISS.

## Author contributions

Conceptualization: RL, MG, NA, IM

Methodology – clinical trials & control samples: RL, MG, KP, JS, BB, DC, TW, NA, IM

Methodology – protein expression: ET, TT, MH, CEC, JH, WHT

Methodology – antibody assays & analytics: RL, KS, RM, NN, TO, MW

Investigation - laboratory: RL, KS, SH, RM

Investigation – analytics: RL, KS, TO, MW

Visualization: RL, KS, TO, MW

Supervision: NA, IM

Writing – original draft: RL

Writing – reviewing & editing: RL, MG, KS, SH, KP, NN, TO, RM, ET, TT, MH, CEC, DC, KT, CD, JH, WHT, JS, MW, BB, TW, NA, TM

## Data and materials availability

All data are available in the main text or the supplementary materials. Additional File 1 contains patient variables, Additional File 2 contains IgG antibody data from the *P. knowlesi* cohorts and Additional File 3 IgG antibody data from the malaria-naïve controls.

## Supplementary Information

**Figure S1:**
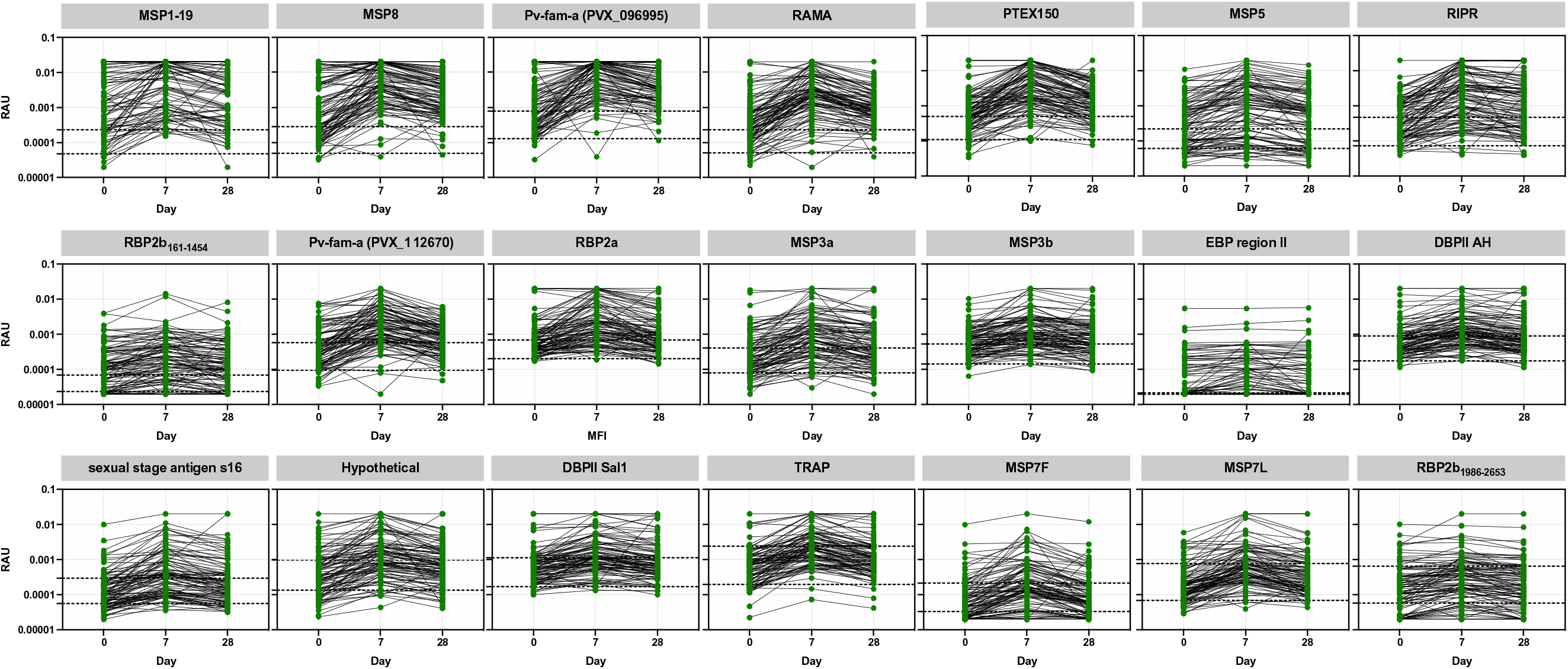
IgG antibody levels against 21 *P. vivax* proteins in 98 individuals with matched data over time. IgG levels were measured against the 21 *P. vivax* proteins using a multiplexed antibody assay. Samples were obtained and run at the time of *P. knowlesi* infection (day 0), and days 7 and 28 following enrolment (ACTKNOW cohort). Results are expressed as the relative antibody units (RAU). Proteins are ordered by highest level of median IgG at day 7 compared to the seropositivity cut-off. Dashed lines indicate the malaria-naïve negative control samples: lower = average of the negative control samples; upper = seropositivity cut-off (average plus 2x standard deviation).

**Figure S2:**
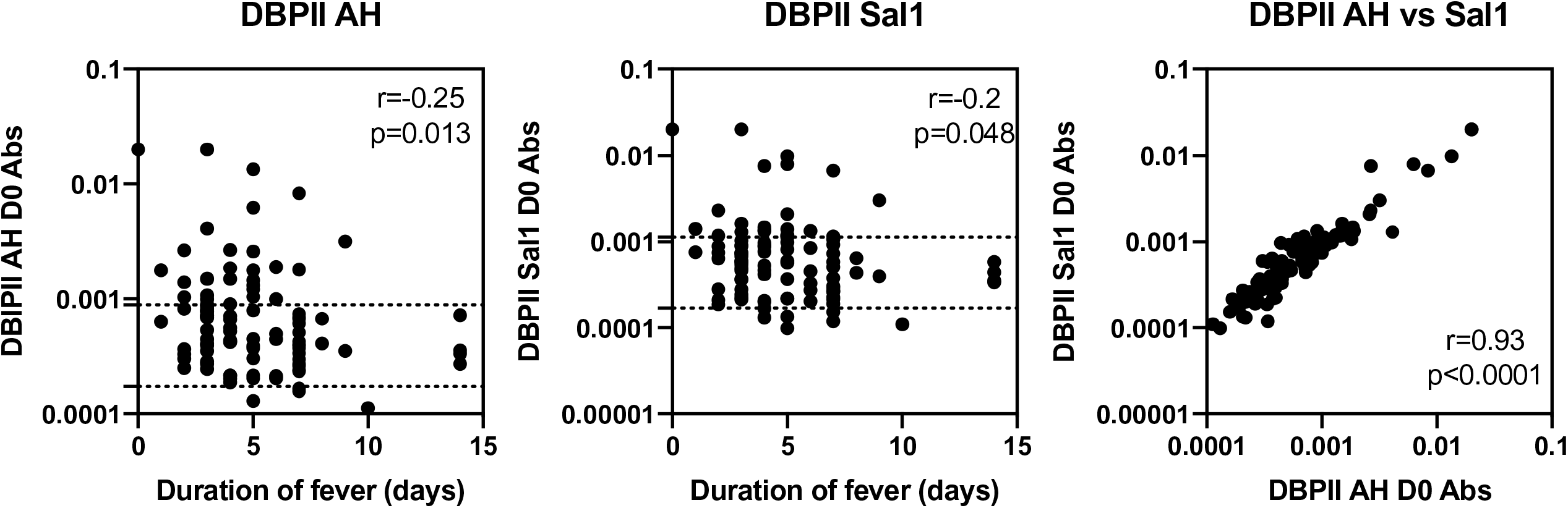
Correlation between acquisition of IgG antibodies at Day 0 to *P. vivax* DBPII constructs and duration of fever of *P. knowlesi* patients. IgG levels were measured against the 21 *P. vivax* proteins using a multiplexed antibody assay. Samples were obtained and run at the time of *P. knowlesi* infection (day 0), and days 7 and 28 following enrolment (ACTKNOW cohort). Results are expressed as the relative antibody units (RAU). Here, IgG antibody data at day 0 were correlated with the duration of fever (days) reported by the *P. knowlesi* patients. Data are shown for the only two proteins with a statistically significant correlation, DBPII AH and Sal1. Dashed lines indicate the malaria-naïve negative control samples: lower = average of the negative control samples; upper = seropositivity cut-off (average plus 2x standard deviation). The third plot shows the correlation between antibodies between both DBPII constructs. Spearman correlation co-efficients are shown.

**Figure S3:**
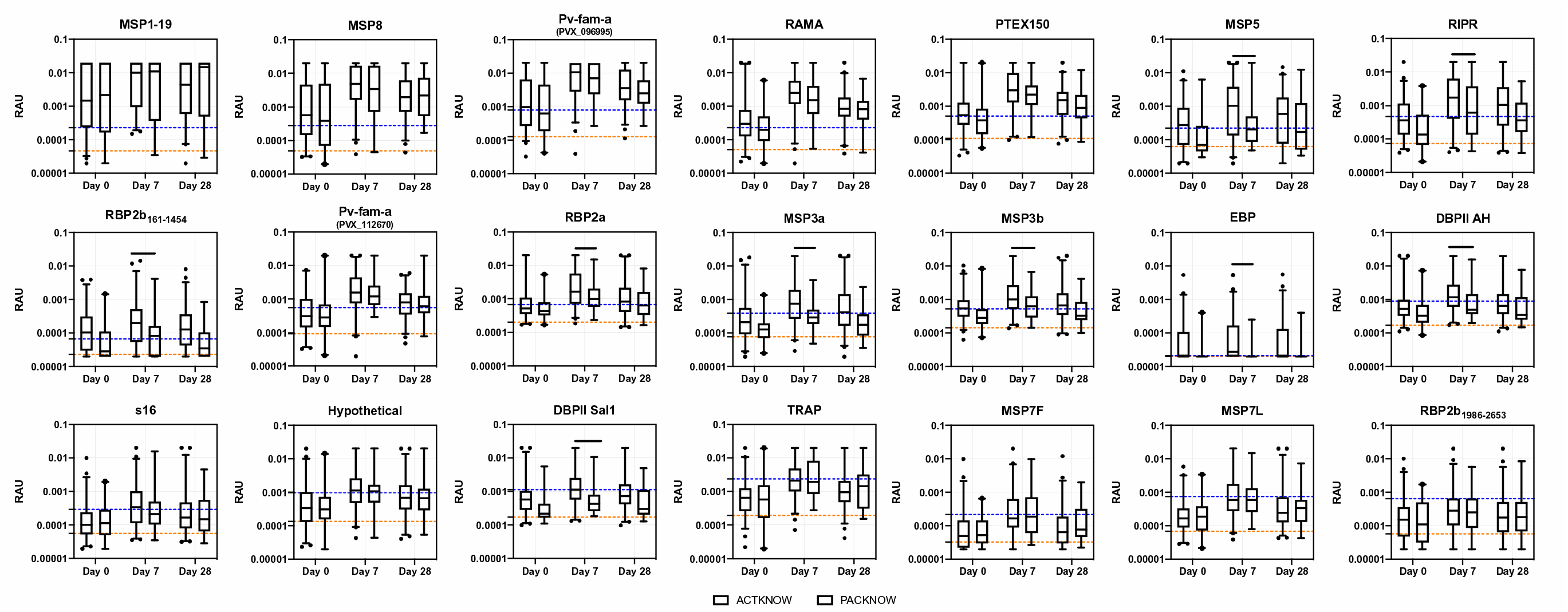
IgG antibody levels against 21 *P. vivax* proteins in *P. knowlesi* patients split via clinical trial. IgG levels were measured against the 21 *P. vivax* proteins using a multiplexed antibody assay. Samples were obtained and run at the time of *P. knowlesi* infection (day 0), and days 7 and 28 following enrolment (ACTKNOW and PACKNOW cohorts). Further data timepoints were available for PACKNOW but are not shown. Results are expressed as the relative antibody units (RAU). Proteins are ordered by highest level of median IgG at day 7 compared to the seropositivity cut-off. Dashed lines indicate the malaria-naïve negative control samples: lower = average of the negative control samples; upper = seropositivity cut-off (average plus 2x standard deviation). Data are split via clinical trial; the ACTKNOW cohort (n=99) and the PACKNOW cohort (n=41 at day 0). Statistical difference in antibody levels between clinical trial groups was only assessed at the peak timepoint of day 7, using Mann-Whitney U tests. 9 of 21 *P. vivax* proteins induced significantly higher levels in ACTKNOW vs PACKNOW: RBP2b_161-1454_ (p=0.0019), MSP3b (p=0.0095), DBPII AH (p=0.0064), RIPR (p=0.013), MSP3a (p=0.0002), MSP5 (p=0.021), RBP2a (p=0.046), DBPII Sal1 (p=0.0049) and EBP (p=0.0007).

**Table S1:** Proteins used in this study.

| Protein | PlasmoDB code/GenBank ID | Full Annotation | Region, amino acids (size) | Expression system | µg protein coupled to 2.5x10<sup>6</sup> beads |
| --- | --- | --- | --- | --- | --- |
| RBP2b<sub>161-1454</sub> | PVX_094255 | Reticulocyte binding protein 2b (n-terminal fragment) | 161-1454 (1294) | *E. coli*, WEHI | 0.6 |
| MSP1-19 | PVX_099980 | Merozoite surface protein 1 - 19 | 1622-1729 (108) | WGCF, CellFree Sciences | 4 |
| RBP2b<sub>1986-2653</sub> | PVX_094255 | Reticulocyte binding protein 2b (c-terminal fragment) | 1986-2653 (667) | WGCF, CellFree Sciences | 8 |
| EBP | KMZ83376.1 | Erythrocyte binding protein | 109-432 (324) | *E. coli*, Institut Pasteur | 0.5 |
| s16 | PVX_000930 | Sexual stage antigen S16 | 31-end (110) | WGCF, CellFree Sciences | 2 |
| RIPR | PVX_095055 | Rh5 interacting protein | 552-1075 (524) | *E. coli*, WEHI | 2 |
| MSP3a | PVX_097720 | Merozoite surface protein 3 | 25-end (828) | WGCF, CellFree Sciences | 2 |
| Hypothetical | PVX_097715 | Hypothetical protein | 20-end (431) | WGCF, CellFree Sciences | 3 |
| DBPII AH | AAY34130.1 | Duffy binding protein region II (strain AH) | 1-237 (237) | *E. coli*, Institut Pasteur | 1.4 |
| MSP8 | PVX_097625 | Merozoite surface protein 8 | 24-463 (440) | WGCF, CellFree Sciences | 1.4 |
| Pv-fam-a | PVX_112670 | Pv-fam-a | 34-end (302) | WGCF, CellFree Sciences | 2.26 |
| RAMA | PVX_087885 | Rhoptry associated membrane antigen | 462-730 (269) | WGCF, CellFree Sciences | 1.2 |
| Pv-fam-a | PVX_096995 | Pv-fam-a | 61-end (420) | WGCF, CellFree Sciences | 3 |
| MSP3b | PVX_097680 | Merozoite surface protein 3 | 21-end (996) | WGCF, CellFree Sciences | 0.8 |
| MSP7L | PVX_082700 | Merozoite surface protein 7 | 23-end (397) | WGCF, CellFree Sciences | 1.5 |
| MSP5 | PVX_003770 | Merozoite surface protein 5 | 23-365 (343) | WGCF, CellFree Sciences | 0.2 |
| MSP7F | PVX_082670 | Merozoite surface protein 7 | 24-end (388) | WGCF, CellFree Sciences | 1 |
| TRAP | PVX_082735 | Thrombospondin related adhesion protein (also known as SSP2) | 26-493 (468) | WGCF, CellFree Sciences | 2 |
| DBPII Sal1 | PVX_110810 | Duffy binding protein region II (strain Sal1) | 193-521 (329) | *E. coli*, Institut Pasteur | 1.5 |
| PTEX150 | PVX_084720 | translocon component PTEX150, putative | 24-908 (885) | WGCF, CellFree Sciences | 1 |
| RBP2a | PVX_121920 | Reticulocyte binding protein 2a | 160-1135 (976) | *E. coli*, WEHI | 1.5 |

**Table S2:** Association between peak antibody levels at day 7 with parasitaemia and sex. Sample size = 134. Regression analyses were performed univariably. Antigens are ordered by the fold change in the peak antibody level at day 7 compared to the seropositivity cut-off based on the negative control samples (=Fold Δ IgG). Fold change data shown is from the ACTKNOW cohort. CI = confidence interval.

| Protein | Fold $\Delta$ IgG | Parasitaemia | | Sex | |
| --- | --- | --- | --- | --- | --- |
|  |  | Coefficient (95% CI) | p value | Coefficient (95% CI) | p value |
| MSP1-19 | 43.98 | -2.22e-6 (-8.69e-6-4.25e-6) | 0.498 | 0.051 (-0.23-0.33) | 0.724 |
| MSP8 | 17.07 | -3.31e-7 (-3.9e-6 - 3.24e-6) | 0.855 | 0.17 (-0.06-0.42) | 0.146 |
| Pv-fam-a (PVX_096995) | 13.39 | -5.35e-7 (-4.02e-6-2.94e-6) | 0.761 | 0.12 (-0.07-0.32) | 0.207 |
| RAMA | 10.86 | -1.41e-6 (-4.02e-6-1.2e-6) | 0.287 | 0.23 (-0.015-0.47) | 0.065 |
| PTEX150 | 6.08 | 3.64e-7 (-2.59e-6-3.32e-6) | 0.808 | 0.11 (-0.13-0.34) | 0.376 |
| MSP5 | 4.70 | -1.92e-6 (-5.28e-6-1.43e-6) | 0.258 | 0.34 (0.049-0.64) | 0.023 |
| RIPR | 3.66 | -2.8e-6 (-7.78e-6-2.18e-6) | 0.267 | 0.25 (-0.015-0.51) | 0.065 |
| RBP2b<sub>161-1454</sub> | 3.00 | 1.59e-6 (-3.74e-6 - 6.95e-6) | 0.557 | 0.073 (-0.18-0.32) | 0.56 |
| Pv-fam-a (PVX_112670) | 2.76 | 9.23e-7 (-1.62e-6-3.46e-6) | 0.473 | -0.036 (-.24-0.17) | 0.724 |
| RBP2a | 2.45 | -2.78e-6 (-5.99e-6-4.35e-7) | 0.09 | -0.02 (-2.2-0.18) | 0.842 |
| MSP3b | 1.93 | 7.7e-7 (-1.16e-6 - 2.7e-6) | 0.432 | 0.26 (0.053-0.46) | 0.014 |
| MSP3a | 1.92 | -7.73e-7 (-3.5e-6-1.95e-6) | 0.575 | 0.28 (0.047-0.52) | 0.019 |
| EBP | 1.33 | -2.02e-6 (-4.83e-6-7.94e-7) | 0.158 | 0.057 (-0.16-0.27) | 0.603 |
| DBPII AH | 1.31 | -3.6e-9 (-5.52e-6 - -6.03e-7) | 0.015 | 0.178 (0.005-0.35) | 0.044 |
| S16 | 1.18 | 8.4e-7 (-3.57e-6-5.25e-6) | 0.707 | 0.23 (-0.035-0.49) | 0.089 |
| Hypothetical | 1.16 | -7.69e-7 (-4.23e-6-2.69e-6) | 0.661 | 0.18 (-0.067-0.430) | 0.152 |
| DBPII Sal1 | 1.02 | -2.13e-6 (-5.29e-6-1.03e-6) | 0.185 | 0.17 (-0.022-0.36) | 0.082 |
| TRAP | 0.90 | 3.31e-7 (-3.95e-6 - 4.61e-6) | 0.878 | 0.11 (-0.096-0.31) | 0.297 |
| MSP7L | 0.78 | -8.79e-7 (-3.09e-6-1.34e-6) | 0.434 | 0.16 (-0.077-0.40) | 0.185 |
| MSP7F | 0.78 | 4.55e-6 (1.29e-6-7.82e-6) | 0.007 | 0.16 (-0.093-0.41) | 0.217 |
| RBP2b<sub>1986-2653</sub> | 0.44 | 2.67e-6 (-1.06e-6 - 6.4e-6) | 0.159 | 0.20 (-0.055-0.45) | 0.125 |

